# Longitudinal Proteogenomic Analysis Reveals Mechanistic Insights into the Progression from Prediabetes to Type 2 Diabetes

**DOI:** 10.64898/2026.02.13.26346161

**Authors:** Archit Singh, Marlene Ganslmeier, Mauro Tutino, Young-Chan Park, Jürgen Machann, Fritz Schick, Andreas Peter, Rainer Lehmann, Yiying Wang, Yurong Cheng, Leontine Sandforth, Sebastian Schuth, Jochen Seissler, Nikolaos Perakakis, Peter E. H. Schwarz, Julia Szendrödi, Robert Wagner, Michele Solimena, Annette Schürmann, Stefan Kabisch, Andreas F. H. Pfeiffer, Stefan R. Bornstein, Matthias Blüher, Norbert Stefan, Andreas Fritsche, Hubert Preissl, Reiner Jumpertz von Schwartzenberg, Martin Hrabe de Angelis, Michael Roden, Ozvan Bocher, Eleftheria Zeggini, Andreas L. Birkenfeld

**Affiliations:** Institute of Translational Genomics, Helmholtz Zentrum München- German Research Center for Environmental Health, Neuherberg, 85764, Germany; Munich School for Data Science (MUDS), Helmholtz Zentrum München- German Research Center for Environmental Health, Neuherberg, 85764, Germany; Doctoral program of Experimental Medicine and Health Sciences, TUM School of Medicine and Health, Technical University of Munich, Munich, 81675, Germany; Institute for Diabetes Research and Metabolic Diseases of the Helmholtz Center Munich at the University of Tübingen, Tübingen, Germany; German Center for Diabetes Research (DZD E.V.), Neuherberg, Germany; Department of Radiology, Section on Experimental Radiology, University of Tübingen, Otfried-Müller-Str. 51, 72076 Tübingen, Germany; Institute for Clinical Chemistry and Pathobiochemistry, Department for Diagnostic Laboratory Medicine, University Hospital of Tübingen, Hoppe-Seyler-Str. 3, 72076 Tübingen, Germany; Department of Diabetology, Endocrinology, Nephrology, University of Tübingen, Tübingen, Germany; Diabetes Research Group, Medical Department 4, Ludwig-Maximilians University Munich, Ziemssenstr. 1, 80336 Munich, Germany; Department of Internal Medicine III, Technical University Dresden, Fetscherstrasse 74, 01307 Dresden, Germany; Department of Medicine I and Clinical Chemistry, University Hospital of Heidelberg, Im Neuenheimer Feld 410, 69120 Heidelberg, Germany; Department of Endocrinology and Diabetology, Medical Faculty and University Hospital, Heinrich Heine University Düsseldorf, Moorenstr. 5, 40225 Düsseldorf, Germany; Institute for Clinical Diabetology, German Diabetes Center, Leibniz Center for Diabetes Research at Heinrich Heine University Düsseldorf, Auf’m Hennekamp 65, 40225 Düsseldorf, Germany; Paul-Langerhans-Institut Dresden (PLID) of the Helmholtz Center Munich at University Clinic Carl Gustav Carus, TU Dresden. Tatzberg 47/49, 01307 Dresden, Germany; Charité University Hospital Berlin; Clinic of Endocrinology and Metabolism, Campus Benjamin Franklin, Hindenburgdamm 30, 12203 Berlin, Germany; Department of Medicine III, University Hospital Carl Gustav Carus, Dresden, Germany; Department of Diabetes, School of Life Course Science and Medicine, King’s College London, London, UK; Department of Endocrinology, Diabetology and Clinical Nutrition, University Hospital Zurich, Zurich, Switzerland; Institute of Experimental Genetics, IEG Helmholtz Center Munich, Ingolstädter Landstraße 1, 85764, Neuherberg, Germany; Chair of Experimental Genetics, School of Life Sciences Weihenstephan, Technical University of Munich, Arcisstraße 21, 80333 Munich, Germany; Univ Brest, Inserm, EFS, UMR 1078, Brest, France; Technical University of Munich (TUM), TUM University Hospital, TUM School of Medicine and Health, 81675 Munich, Germany

**Author notes:** These authors are joint first authors (Archit Singh, Marlene Ganslmeier). These authors are joint last authors (Martin Hrabe de Angelis, Michael Roden, Ozvan Bocher, Eleftheria Zeggini, Andreas L. Birkenfeld).

## Abstract

Prediabetes and type 2 diabetes (T2D) are metabolic disorders characterized by insulin resistance and β-cell dysfunction. To understand the molecular mechanisms driving the transition from prediabetes to T2D, we performed a longitudinal proteogenomic analysis on 458 participants from the Prediabetes Lifestyle Intervention Study (PLIS). We identified 185 plasma proteins to be differentially expressed between conditions, 36 of which predict future T2D-onset. Integrating genetic data from 321 individuals, we generated a genome-wide protein quantitative trait loci (pQTL) map, identifying 86 differential and 700 shared *cis*-pQTLs between prediabetes and T2D. Mediation analysis revealed 60 putative causal links connecting allele-driven plasma protein expression to clinical traits, identifying body fat distribution, insulin resistance, and β-cell function as central drivers of pathogenesis. Collectively, these findings highlight specific proteins underlying disease progression and substantiate the view that prediabetes and T2D are not distinct conditions, but rather stages on a unified metabolic spectrum.

## Introduction

T2D is characterized by chronic hyperglycemia and ranks among the most consequential drivers of global disease burden [1]. Prediabetes is recognized as an early stage in the continuum of glucose dysregulation, whereas T2D reflects a more advanced manifestation of the same disorder [2]. Compared with the normoglycemic stage, both conditions share core abnormalities, underscoring prediabetes as a key risk state for subsequent T2D. Progression from prediabetes to overt T2D is largely driven by worsening insulin resistance coupled with a gradual decline in pancreatic β-cell function [3]. While recent studies have improved our understanding of risk factors associated with this transition, the mechanisms that govern progression remain incompletely defined, and there is still a need for early, robust biomarkers that reliably capture—and ideally predict—the evolution from normoglycemia to prediabetes to T2D [3–6].

Omics technologies have greatly advanced our understanding of T2D and its underlying mechanisms. With declining costs, these approaches are increasingly being deployed to interrogate metabolic disorders at scale. In particular, recent plasma proteomic studies in T2D have provided valuable insights into disease biology and have nominated candidate biomarkers [7–9].

By contrast, shifts in plasma protein profiles across the transition from prediabetes to overt T2D have been far less well characterized. To date, longitudinal data are largely lacking, and only a limited number of cross-sectional analyses have compared plasma proteomes between prediabetes and T2D [6, 8]. These studies reported multiple proteins associated with both prediabetes and T2D, as well as with longitudinal changes in related clinical traits [8]. In a complementary short-term longitudinal study, circulating protein biomarkers were identified in individuals with recent-onset T2D and linked to insulin resistance, obesity, hyperglycemia and liver steatosis [7]. The paucity of structured, longer term longitudinal investigations has constrained mechanistic insight into the pathophysiology of progression from prediabetes to T2D. Yet such studies are essential to reveal the biological processes that drive disease development and to establish predictive biomarkers for risk stratification. Therefore, we sought to define plasma proteome of progression from prediabetes to T2D, with the goal of gaining deeper insight into mechanisms underpinning disease progression/non-progression and/or persistence.

Plasma protein abundances are, in part, under genetic control through pQTLs. Mapping pQTLs provides a powerful framework to quantify how inherited genetic variation shapes circulating protein levels and, in turn, to illuminate biological pathways and mechanisms implicated in health and disease [10]. Integrating proteomics with genomics shows that genetically regulated proteins contribute to T2D-related traits [11]. Comparing pQTL patterns in prediabetes versus T2D may reveal genetic underpinnings of early molecular changes linked to progression or maintenance and help prioritize biomarkers and therapeutic targets.

Here, we report a longitudinal proteogenomic analysis of individuals with existing prediabetes from the randomized, multicenter PLIS cohort, conducted in nine hospitals in the framework of the German Center for Diabetes Research (DZD). Participants completed a 1-year lifestyle intervention and were followed for 5 years thereafter (total 6-year follow-up) [4, 12]. We quantified ∼3,000 plasma proteins to assess long-term proteomic changes and associations with metabolic phenotypes, including body fat distribution comprised of visceral adipose tissue (VAT) volume and intrahepatic lipid (IHL) content assessed by MRI/^1^H-MRS, and T2D-related glucose traits, including repeated 5-point oral glucose tolerance testing (OGTT). We then mapped pQTLs in individuals who remained with prediabetes and those who progressed to T2D at follow-up, performed differential pQTL analyses to define shared versus condition-specific genetic regulation, and applied causal inference to prioritize plasma proteins linked to T2D-related traits. Collectively, these data identify key biomarkers of T2D progression, provide mechanistic insights into pathogenesis, and characterize the overall proteogenomic signature of the transition from prediabetes to T2D.

## Results

### PLIS Cohort Characteristics

A total of 458 participants from PLIS were included for which genotyping, plasma proteome profiling and deep phenotyping were available at baseline, and at the 6-year follow-up timepoint. The basis for inclusion and exclusion of patients is given in the CONSORT flow chart (Supplementary Figure 1). Baseline characteristics of the whole group are given in Supplementary Table 1. We further categorized participants by glycemic status at the 6-year follow-up time point into normal glucose regulation (NGR), prediabetes (according to ADA criteria) or T2D (according to ADA criteria). Table 1 summarizes baseline characteristics of participants with prediabetes, stratified by glycemic status at the 6-year follow-up into NGR, prediabetes, or T2D.

**Table 1:** Baseline and 6-year follow-up characteristics of the PLIS cohort. All participants had prediabetes at baseline and are shown here stratified by the glucose regulation category they were assigned to at follow-up. NGR = Normal Glucose Regulation, PreD = Prediabetes, T2D = Type 2 Diabetes at follow-up. Between group comparisons of anthropometric and clinical traits at baseline and follow-up have been shown with significant differences highlighted in bold. BMI = Body Mass Index, VAT [L] = Visceral Adipose Tissue [Liters], IHL = Intrahepatic Lipids, FLI = Fatty Liver Index, HOMA-IR = Homeostasis Model Assessment for Insulin Resistance, OGIS = Oral Glucose Insulin Sensitivity Note that Disposition index measures the pancreatic β-cell function and the Insulinogenic index measures how effectively the pancreas releases insulin in the early phase after eating.

| Variable | Baseline |  |  |  | Follow-up |  |  |  |
| --- | --- | --- | --- | --- | --- | --- | --- | --- |
|  | NGR<br>N = 49<sup>1</sup> | PreD<br>N = 321<sup>1</sup> | T2D<br>N = 88<sup>1</sup> | p-value<sup>2</sup> | NGR<br>N = 49<sup>1</sup> | PreD<br>N = 321<sup>1</sup> | T2D<br>N = 88<sup>1</sup> | p-value<sup>2</sup> |
| Sex |  |  |  | 0.7 |  |  |  | 0.7 |
| Female | 24 (49%) | 174 (54%) | 49 (56%) |  | 24 (49%) | 174 (54%) | 49 (56%) |  |
| Male | 25 (51%) | 147 (46%) | 39 (44%) |  | 25 (51%) | 147 (46%) | 39 (44%) |  |
| Age | 53 [46-64] | 62 [55-68] | 62 [54-67] | <0.001 | 60 [52-71] | 68 [62-74] | 68 [60-73] | <0.001 |
| BMI | 29.9 [26.9-33.6] | 28.7 [25.8-32.3] | 30.1 [27.2-33.9] | 0.042 | 27.6 [25.9-31.9] | 28.3 [25.3-32.1] | 30.5 [27.3-33.8] | 0.004 |
| Body Fat [%] | 34 [24-40] | 33 [26-43] | 35 [27-43] | 0.5 | 32 [22-38] | 31 [24-41] | 37 [26-44] | 0.011 |
| VAT [L] | 4.16 [2.52-7.67] | 4.90 [3.18-6.62] | 5.05 [3.83-6.76] | 0.3 | 3.57 [2.17-6.02] | 4.76 [3.31-6.48] | 5.76 [4.43-6.76] | 0.003 |
| IHL [%] | 5 [2-9] | 5 [2-11] | 7 [5-13] | 0.11 | 2.0 [1.1-3.4] | 3.7 [1.8-10.1] | 12.2 [4.4-15.5] | <0.001 |
| FLI | 65 [34-89] | 63 [35-83] | 73 [52-89] | 0.025 | 41 [18-76] | 54 [26-79] | 79 [54-91] | <0.001 |
| Disposition Index | 2.52 [1.58-3.83] | 2.14 [1.38-3.27] | 1.24 [0.90-2.08] | <0.001 | 2.90 [2.22-4.77] | 2.09 [1.39-3.18] | 0.97 [0.68-1.35] | <0.001 |
| HOMA-IR | 2.79 [2.22-3.95] | 3.03 [2.08-4.25] | 3.37 [2.28-5.11] | 0.13 | 2.34 [1.82-3.30] | 3.22 [2.24-4.71] | 5.17 [3.31-9.02] | <0.001 |
| Insulinogenic Index | 0.95 [0.58-1.61] | 0.81 [0.52-1.51] | 0.62 [0.39-1.23] | 0.003 | 0.96 [0.60-1.54] | 0.81 [0.51-1.26] | 0.57 [0.30-0.89] | <0.001 |
| OGIS | 339 [288-388] | 325 [283-367] | 299 [259-346] | 0.001 | 398 [366-439] | 339 [306-374] | 285 [238-324] | <0.001 |
| Glucose 0 [mg/dL] | 102 [100-106] | 105 [101-111] | 113 [106-119] | <0.001 | 92 [88-97] | 105 [99-111] | 128 [117-136] | <0.001 |
| Glucose 2h [mg/dL] | 123 [102-151] | 132 [110-153] | 151 [132-175] | <0.001 | 103 [86-117] | 127 [106-147] | 214 [183-237] | <0.001 |
| HbA1c [%] | 5.50 [5.30-5.60] | 5.80 [5.60-6.00] | 5.90 [5.70-6.10] | <0.001 | 5.40 [5.20-5.50] | 5.80 [5.60-6.00] | 6.30 [6.00-6.60] | <0.001 |

<sup>1</sup> n (%); Median [Q1-Q3]
<sup>2</sup> Pearson’s Chi-squared test; Kruskal-Wallis rank sum test

At baseline, total body fat content was comparable across all groups. Participants who reverted to NGR at follow-up were significantly younger but otherwise metabolically similar to those who remained in the prediabetic condition (Table 1). In contrast, individuals who progressed to T2D exhibited a significantly higher fatty liver index (FLI), lower disposition index—reflecting impaired pancreatic β-cell function—and reduced insulin sensitivity, along with higher fasting and 2-hour glucose levels compared to individuals who reverted to NGR and remained with prediabetes (Table 1).

At the 6-year follow-up, as expected, participants with T2D showed significantly higher fasting and 2-hour glucose levels as well as higher HbA1c compared with the other groups (Table 1). They also had significantly higher BMI, consistent with increased total body fat, as well as greater VAT volume, IHL content, and FLI (Table 1). These changes were accompanied by significantly higher insulin resistance and reduced insulin secretion (Table 1).

When examining within-group changes over time, individuals with NGR and prediabetes exhibited significant improvements in body fat %, VAT volume, IHL content, FLI, along with a significant increase in insulin sensitivity at the 6-year follow-up compared to baseline (Supplementary Table 2). In contrast, individuals who progressed to T2D demonstrated a significant deterioration in key clinical features, including increased VAT volume, impaired pancreatic β-cell function, heightened insulin resistance, and reduced insulin sensitivity relative to baseline.

### Baseline plasma proteomics show minimal differences across future outcome groups, but longitudinal changes emerge with progression to T2D

To determine whether the baseline plasma proteomic profile could predict glycemic outcomes at follow-up, we compared baseline protein expression between individuals who achieved remission to NGR and those who remained in the prediabetic condition 6-years later. Here we did not observe significant differences between the two groups (Supplementary Figure 2A). Similarly, baseline plasma protein expression did not differ significantly between participants who progressed to T2D and those who remained in the prediabetic condition (Supplementary Figure 2B).

We next evaluated whether longitudinal changes in plasma protein expression reflected changes in glycemic status by comparing baseline versus follow-up within each outcome group. No significant within-group changes were observed in the NGR remission group (Supplementary Figure 3A). In the persistent prediabetes group, 12 proteins changed significantly between baseline and follow-up (Supplementary Figure 3B), including lower levels of C1QL2, CXCL5, IL7, and CCL5 at follow-up; these proteins are implicated in inflammatory and immune signaling pathways. These findings may be explained by the lifestyle intervention, which leads to rather improved metabolic regulation within the prediabetes stratum, as highlighted in the previous section and Supplementary Table 2.

In the group progressing from prediabetes to T2D, CASP4 increased at follow-up relative to baseline (Supplementary Figure 3C); CASP4 is a caspase-family protein involved in apoptosis and inflammatory responses [13].

### Cross-sectional follow-up plasma proteomics identifies an emergent signature of progression to T2D

As there were no proteomic differences at baseline—when all participants lived with prediabetes (Supplementary Figure 2)—any proteomic differences observed at the 6-year follow-up must have emerged during the follow-up period. To identify these cross-sectional differences at follow-up, we compared the outcome groups. Plasma protein expression did not differ between individuals in remission to NGR and those with persistent prediabetes (Supplementary Figure 4). In contrast, 185 plasma proteins were differentially expressed between individuals remaining in the prediabetic condition and those advancing to T2D (FDR < 0.05; Supplementary Table 3), with 116 upregulated and 69 downregulated in T2D (Figure 1A). Since these proteins were not differentially expressed at baseline in the same individuals, these findings support the conclusion that the signature develops during progression from prediabetes to T2D. Compared with prior circulating proteomics studies in T2D, 111 of the 185 proteins were novel (Supplementary Tables 3). Gene set enrichment analysis (using GO and KEGG) indicated upregulation of pathways pertaining to lipid and fatty-acid metabolism and branched-chain amino-acid degradation in T2D, whereas apoptosis and autophagy pathways, among others, were downregulated (Figure 1B–C). Previous studies have reported dysregulation of lipid and fatty acid metabolism as well as apoptotic pathways in T2D [14, 15]. Moreover, elevated plasma levels of branched-chain amino acids have been shown to predict future development of T2D [16].

**Figure 1:**
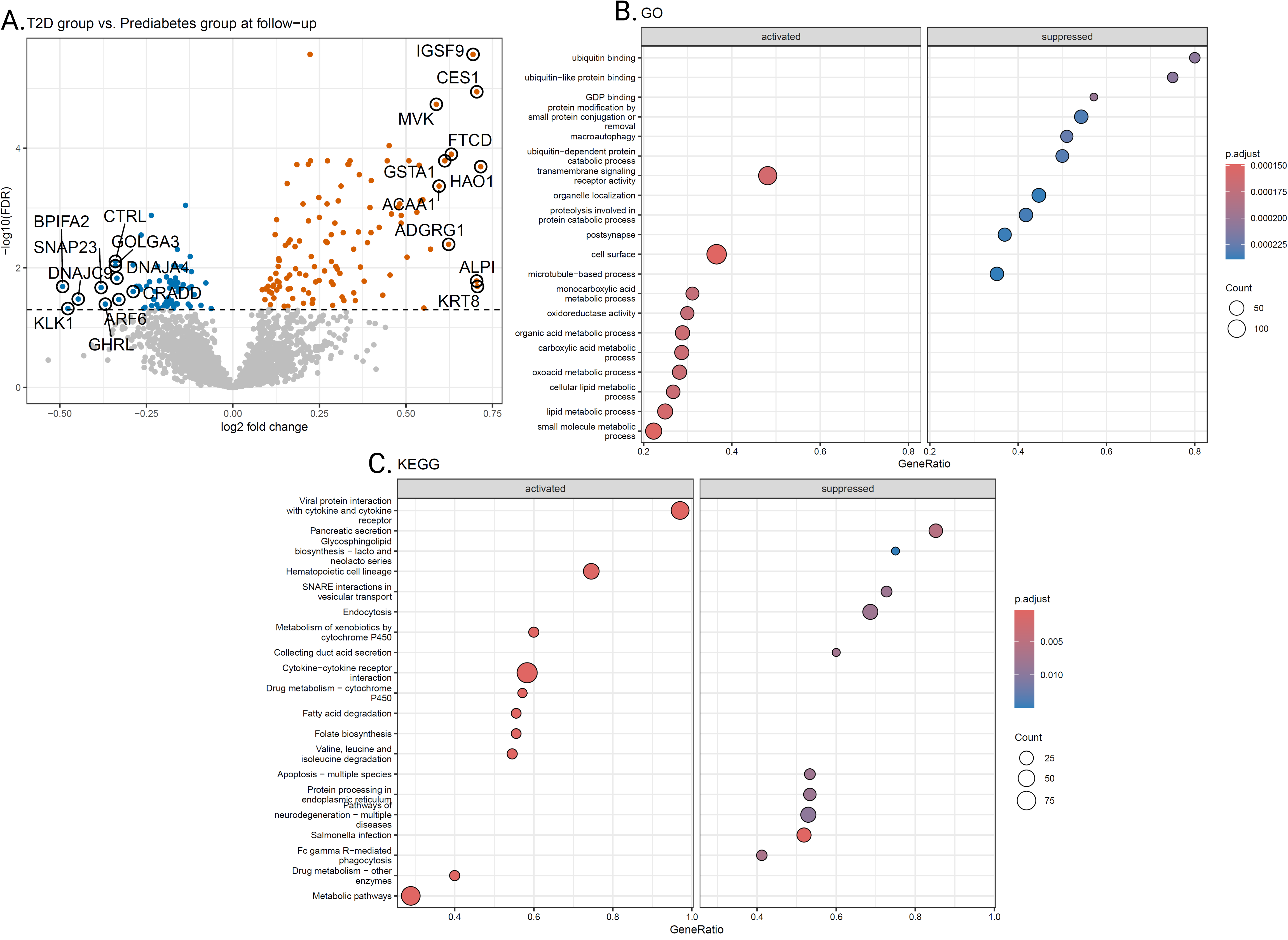
A. Volcano plot showing significantly differentially expressed proteins and highlighting the top 10 upregulated and downregulated proteins in individuals with T2D compared to prediabetes at follow-up. B. Gene set enrichment analysis of significantly differentially expressed proteins using the Gene Ontology database. The plot shows significant upregulation of pathways associated with lipid-related metabolic processes in T2D compared to the prediabetes group at follow-up. C. Gene set enrichment analysis of significantly differentially expressed proteins using the KEGG database. The plot shows significant upregulation of pathways associated with fatty acid metabolism and branched chain amino acid degradation as well as significant downregulation of apoptotic pathways in T2D group compared to the prediabetes group at follow-up.

### Changes in plasma protein expression are associated with T2D development and related clinical features

We next focused on the 185 proteins that were differentially expressed between individuals who remained with prediabetes and those who progressed to T2D at follow-up, as these changes may provide insights into the pathophysiology of diabetes development and overall metabolic decline or may serve as potential biomarkers of decline. To assess whether these protein alterations reflect transient snapshots or dynamic trajectories of disease progression, we examined if longitudinal changes in plasma protein expression were associated with T2D status at follow-up. Of the 185 proteins analyzed, 36 exhibited significant changes in expression association with T2D status at follow-up (Figure 2A), all of which were upregulated in individuals with T2D compared to those with prediabetes. Among these 36 proteins, 11 plasma proteins represent novel potential biomarkers of T2D which are not previously reported in circulating proteome studies of the disease (Supplementary Table 4) and may offer novel insight into disease progression and trajectories [8].

**Figure 2:**
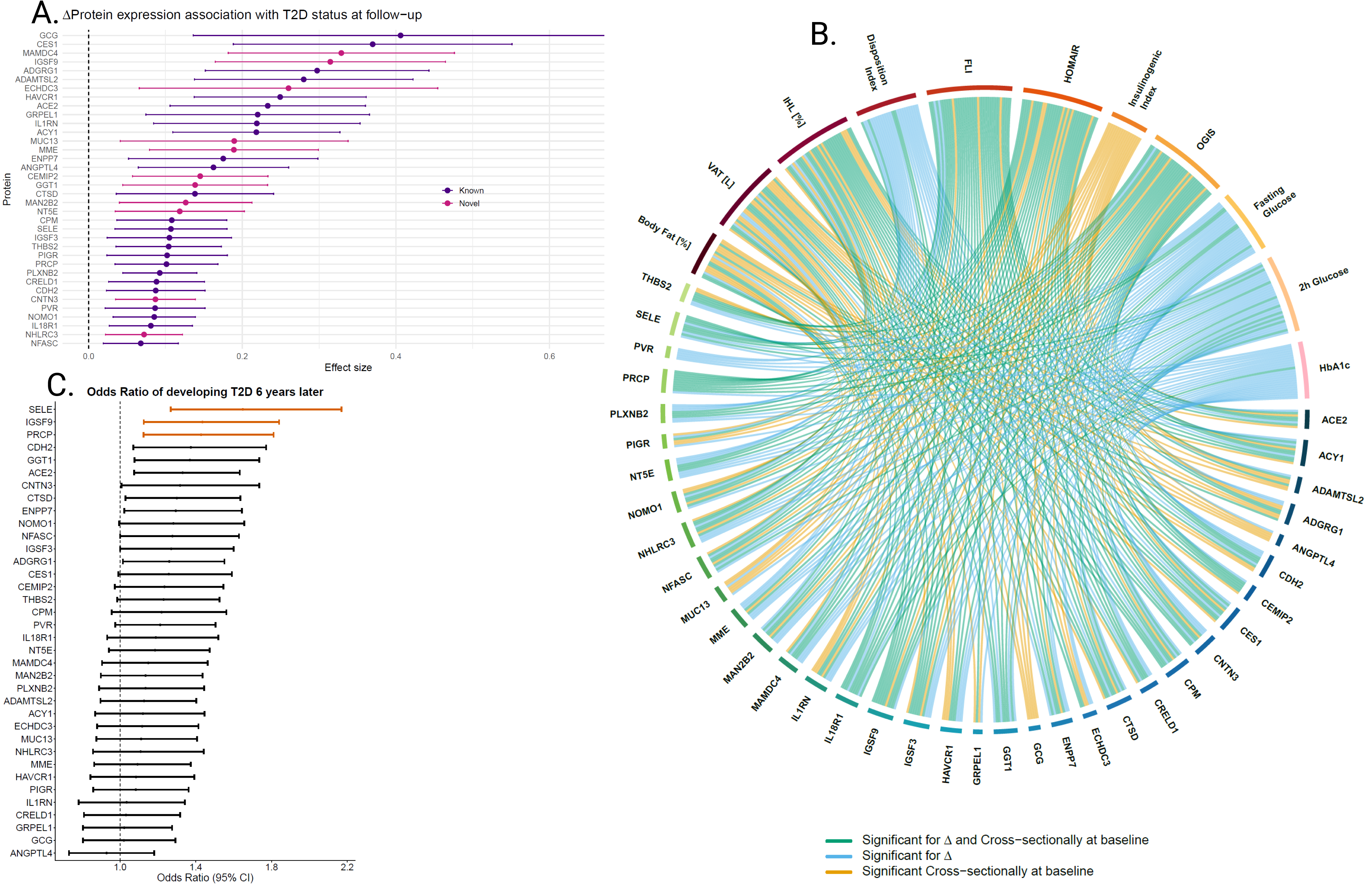
A. Forest plot showing effect sizes of 36 proteins whose change in expression is significantly associated with T2D status at follow-up. The proteins which have not been associated with prediabetes or T2D in previous studies are highlighted in pink. B. Chord plot showing association of 36 plasma proteins with 11 clinical and metabolic features at baseline and longitudinally. The T2D-related features are highlighted in the upper half of the chord plot with different colors while the plasma proteins are highlighted in the lower half of the chord plot with colors ranging between green and blue. VAT [L] = Visceral Adipose Tissue [Liters], IHL = Intrahepatic Lipids, FLI = Fatty Liver Index, HOMA-IR = Homeostasis Model Assessment for Insulin Resistance, OGIS = Oral Glucose Insulin Sensitivity Note that Disposition index measures the pancreatic β-cell function and the Insulinogenic index measures how effectively the pancreas releases insulin in the early phase after eating. C. Forest plot showing the odds-ratios of proteins whose baseline expression is predictive of T2D at follow-up. Among the 36 proteins analyzed, three show significant odds-ratios (FDR < 0.05) highlighted in vermilion.

As T2D is characterized by progressive deterioration of insulin sensitivity and beta-cell function, fasting and 2-hour glucose levels, as well as increased IHL content and VAT volume, we examined associations between 36 proteins and 11 T2D-related clinical phenotypes. These analyses were performed both at baseline and for changes in plasma protein expression and corresponding changes in clinical traits over time (Figure 2B, Supplementary Table 5).

In the baseline analysis, plasma proteins were found to show significant associations with first phase insulin secretion (insulinogenic index), insulin resistance (HOMA-IR), and body composition i.e. VAT volume, and body fat % (Figure 2B, Supplementary Table 5). The strongest positive association was observed between protein interleukin 18 receptor 1 (IL18R1) and HbA1c. Further, the protein glucagon (GCG) showed a negative association with insulin sensitivity, and positive associations with IHL content, FLI, VAT volume and insulin resistance, exclusively at baseline.

Predicting progression to T2D among individuals with prediabetes is of high clinical importance as not all individuals will develop T2D and preventive resources should be prioritized for those at highest risk. In this context, among the 36 proteins assessed at baseline, E-selectin (SELE), Immunoglobulin Superfamily Member 9 (IGSF9), and Prolylcarboxypeptidase (PRCP) were significantly associated with incident T2D at follow-up, showing odds ratios indicative of increased risk of progression (Figure 2C).

In the longitudinal analyses, Δ denotes the within-individual change from baseline to follow-up. Changes in several plasma proteins (Δprotein) were associated with changes in key clinical measure including fasting and 2-hour plasma glucose, HbA1c, insulin sensitivity and resistance, disposition index (β-cell function) and IHL content (Figure 2B; Supplementary Table 5). Among these, IGSF9 showed the strongest positive association with ΔHbA1c and an inverse association with the change in insulin sensitivity. In addition, changes in Poliovirus receptor (PVR) were associated with multiple T2D-related changes including fasting glucose, and 2-hour glucose, insulin sensitivity (OGIS)/ resistance, β-cell function (disposition index) and FLI.

### Genome-wide pQTL mapping identifies genetic determinants of the plasma proteome in prediabetes

To assess the extent to which variation in plasma protein levels is genetically regulated, we performed a genome-wide pQTL analysis of 2,523 plasma proteins in 321 individuals with prediabetes at the follow-up time point. We identified 555 independent pQTLs (Figure 3, Supplementary Table 6) comprising 398 *cis*-associations (71.7%) and 157 *trans*-associations (28.3%) affecting 148 proteins; 18 proteins had both *cis* and *trans* signals. Comparison with 47 published plasma pQTL studies (listed in Supplementary Table 7), revealed 37 novel *trans*-associations, indicating that we determined signals observed in previous studies as well as novel previously unreported signals.

**Figure 3:**
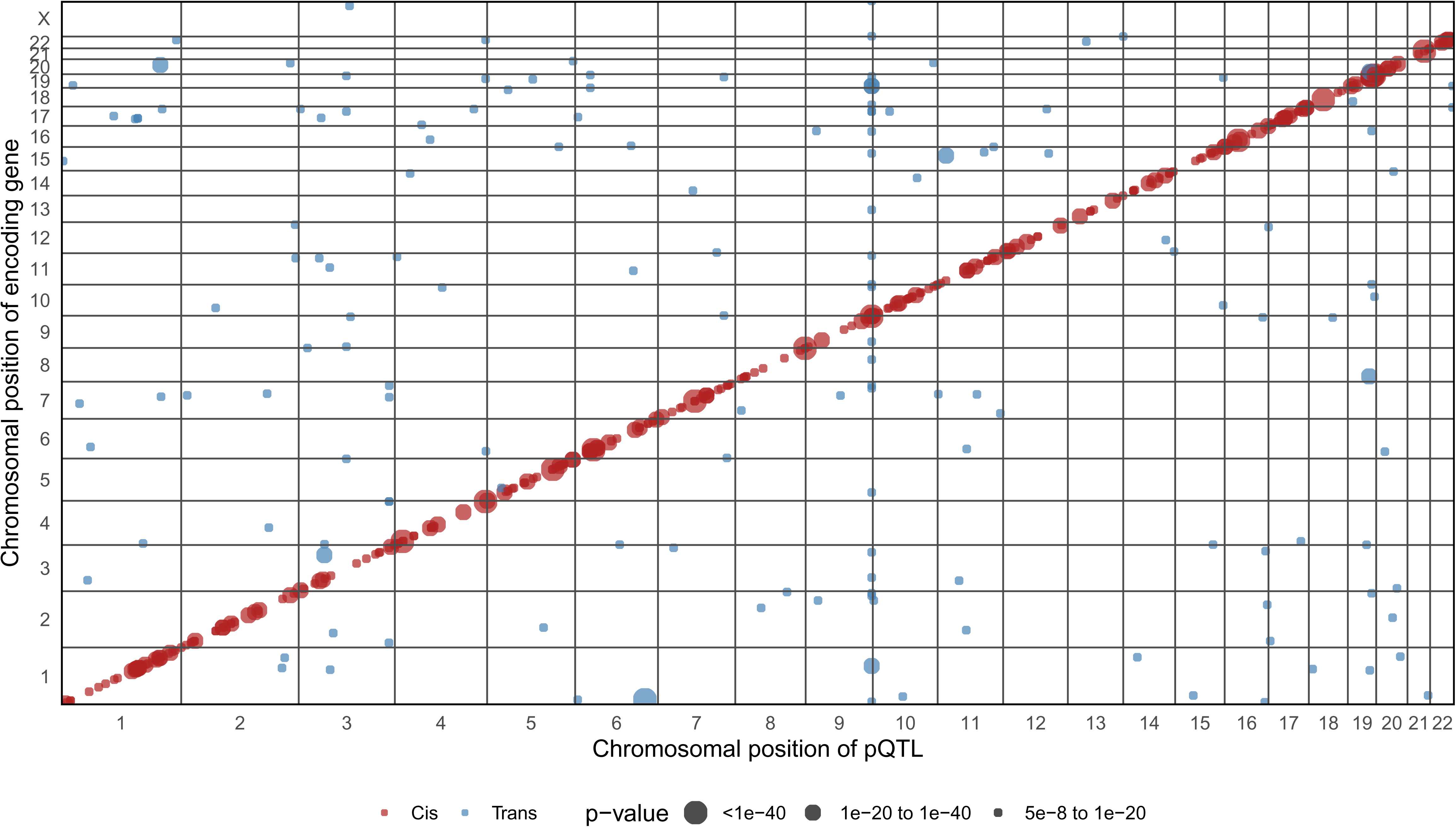
Scatter plot showing the chromosomal positions of independent genome-wide pQTLs and the chromosomal positions of the encoding genes. *Cis*-pQTLs are shown in red and the *trans*-pQTLs are shown in light blue. The dot size refers to the significance of the association.

### Glycaemia-dependent pQTLs implicate stage-specific genetic regulation in progression to T2D

To test whether genetic control of the plasma proteome differs by disease stage—providing a mechanistic link between genotype and progression—we evaluated whether *cis*-pQTL effects change between prediabetes and T2D. Using mash analyses on 2,460 *cis*-pQTLs, we identified 86 variants (3.49%) with significant differential effects in prediabetes relative to T2D (Supplementary Table 8). The strongest association was found for rs6107718 (local false sign rate 3.56×10⁻□), where the G allele was associated with lower Chromogranin B (CHGB) levels specifically in prediabetes (Figure 4A). Notably, three differential pQTLs in prediabetes mapped to proteins also reduced in prediabetes versus T2D— E-selectin (SELE; rs7519364), selenocystein-lyase (SCLY; rs1131296), and tartrate-resistant acid phosphatase type 5 (ACP5; rs12985274) (Figure 4B). These stage-specific genetic effects suggest that part of the proteomic shift observed across glycemic states reflects changes in regulatory architecture, with genetic influences present in prediabetes that are attenuated or absent in established T2D.

**Figure 4:**
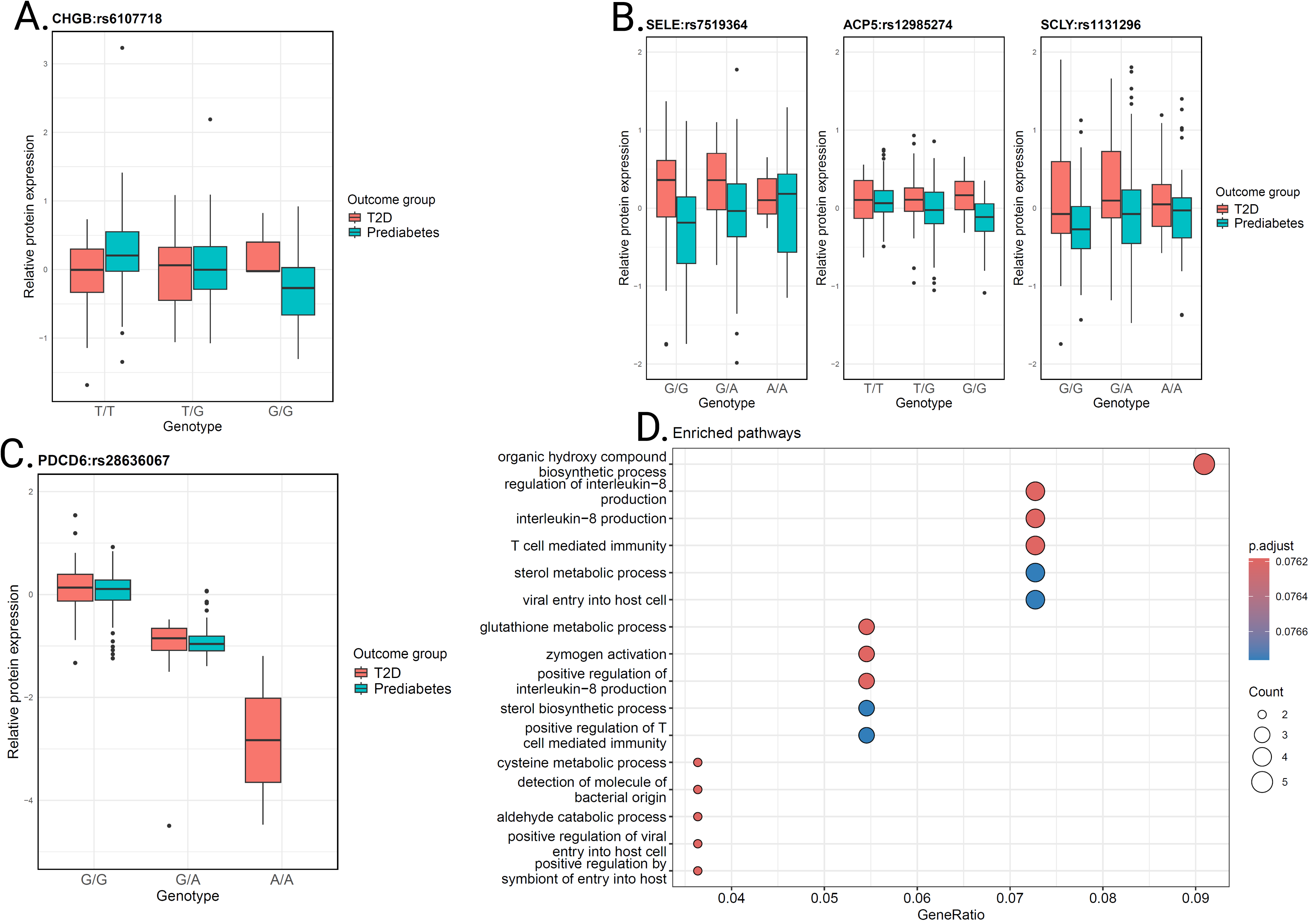
A. Box-whisker plots showing allele dosage of differential pQTL rs6107718 in prediabetes and the relative levels of CHGB protein in individuals. B. Box-whisker plots showing allele dosage of 3 differential pQTLs in prediabetes for proteins SELE, ACP5 and SCLY and their relative levels in individuals. These plasma proteins were found to be significantly downregulated in prediabetes compared to T2D at the follow-up timepoint. C. Box-whisker plots showing allele dosage of shared pQTL rs28636067 between prediabetes and T2D and the relative levels of PDCD6 protein in individuals. D. Pathway over-representation analysis of proteins which were significantly differentially expressed corresponding to shared pQTLs.

Of the 2,460 tested pQTLs, 700 (28.4%) showed shared genetic regulation with concordant effects in both prediabetes and T2D (Supplementary Table 9). The strongest shared signal was rs28636067 (smallest local false sign rate), where the A allele was associated with lower Programmed Cell Death 6 (PDCD6) levels in both conditions (Figure 4C). Notably, 20 shared pQTLs displayed opposite effect directions between prediabetes and T2D (Supplementary Figure 5), suggesting stage-dependent shifts in genetic regulation during disease progression.

Among the 700 proteins with shared pQTLs, 58 were differentially expressed between prediabetes and T2D (46 higher and 12 lower in T2D). Pathway analysis of these 58 proteins implicated IL-8 regulation/production and T-cell–mediated immunity, indicating that immune-related proteomic alterations across glycemic states are partly under stable genetic control (Figure 4D). Sterol metabolism was also enriched, consistent with its reported upregulation in T2D and inverse association with insulin sensitivity [17, 18].

### Causal effects of plasma proteins on clinical features in prediabetes

To explore whether pQTLs may influence clinical traits through plasma protein abundance, we performed mediation analysis. This approach uses allele-associated differences in protein levels to test whether protein abundance is consistent with a mediating role between genotype and phenotype, providing evidence compatible with—but not proving—causal mechanisms.

We tested 86 differential pQTL–protein pairs against 11 clinical features in the prediabetes group related to glucose regulation and body fat distribution and identified 60 significant mediation relationships involving 29 proteins (Figure 5; Supplementary Table 10), each with a significant average causal mediation effect (ACME). Of these, 29 ACMEs were negative, consistent with proteins potentially associated with more favorable trait profiles. The mediated traits were enriched for liver fat (FLI), body fat distribution (body fat %), and insulin sensitivity (OGIS), suggesting that these phenotypes may be particularly connected to genetically influenced variation in plasma proteins during prediabetes and the progression to T2D.

**Figure 5:**
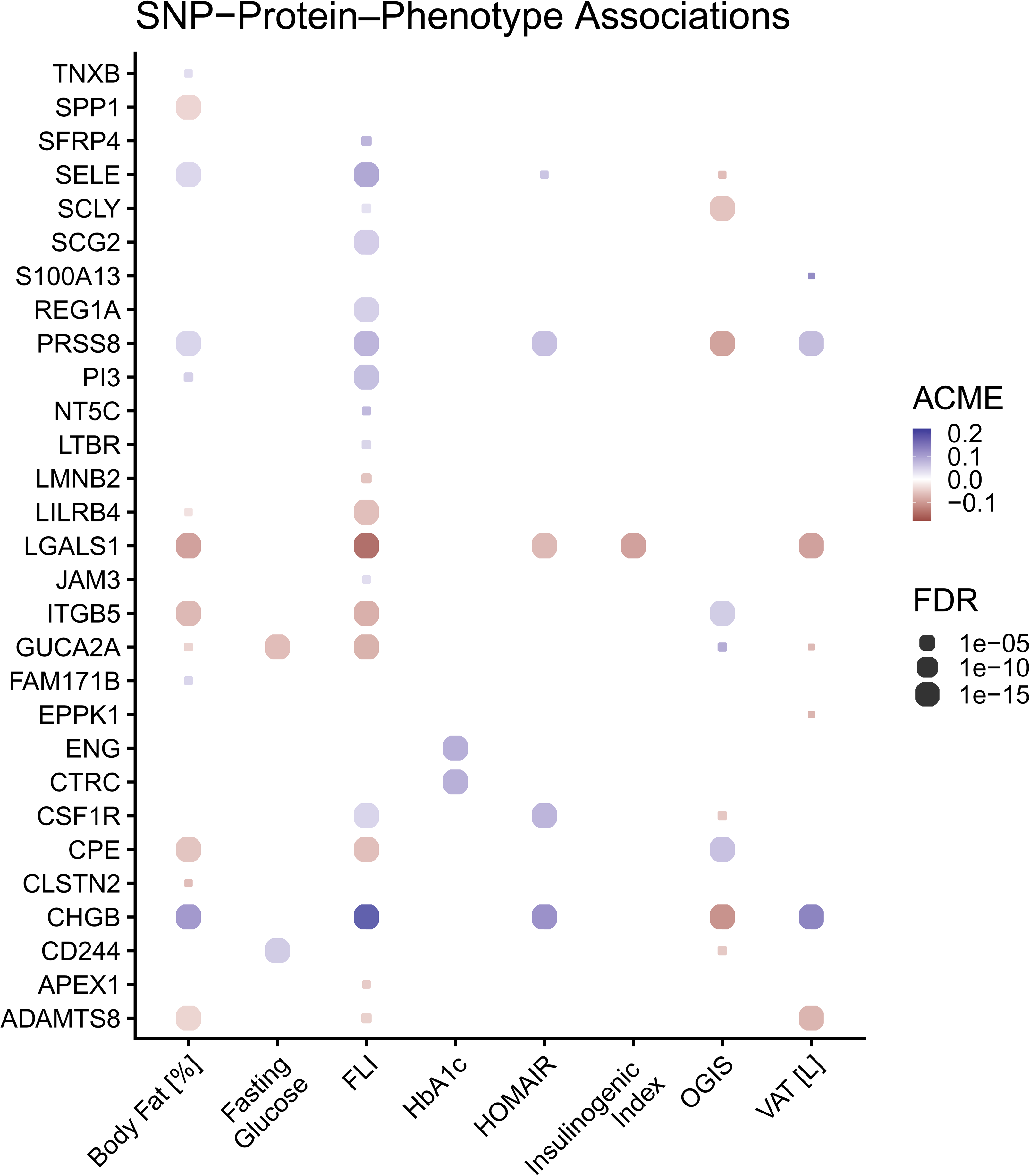
Dot plot showing the results of the mediation analysis. The plot shows proteins with significant ACME after FDR-adjustment with the x-axis showing the clinical features and the y-axis showing the plasma protein corresponding to the differential pQTL. FLI = Fatty Liver Index, HOMA-IR = Homeostasis Model Assessment for Insulin Resistance, OGIS = Oral Glucose Insulin Sensitivity, VAT [L] = Visceral Adipose Tissue [Liters]. Insulinogenic index measures how effectively the pancreas release insulin in the early phase after eating.

## Discussion

In this study, we present the first comprehensive proteogenomic analysis of the transition from prediabetes to T2D, leveraging longitudinal plasma proteomic, genomic and deep phenotypic clinical data from the PLIS randomized controlled clinical study [4, 19]. A major strength is the clinically coherent entry point: all participants had prediabetes, as defined by ADA criteria, at baseline, were deeply phenotyped in a standardized way, including MRI, ^1^H-MRS, 5-point OGTT and harmonized preanalytical procedures, and were followed under a standardized protocol to capture divergent trajectories (persistent prediabetes, remission to NGR, or progression to T2D). This design minimizes heterogeneity at study entry and enables separation of molecular features that precede progression from those that emerge with worsening glycaemia.

Two findings provide valuable insights into the temporal dynamics of the plasma proteome across metabolic disease stages. At baseline, when all individuals were still prediabetic, baseline plasma proteomic profiles did not differ between those who later achieved remission to NGR, remained prediabetic, or progressed to T2D, indicating that broad proteomic separation is not readily detectable at the prediabetic stage (Supplementary Figure 2). In contrast, cross-sectional comparisons at the 6-year follow-up revealed a pronounced proteomic signature of established T2D relative to prediabetes with 185 differentially expressed plasma proteins. The absence of baseline differences, together with the emergence of strong between-group differences at follow-up, supports the interpretation that proteomic divergence largely unmasks during progression to overt T2D rather than reflecting stable pre-existing differences. This finding further strengthens the view that prediabetes and T2D lie on a continuum within a prolonged trajectory of glucose dysregulation, in which molecular alterations accumulate and become more pronounced as metabolic control deteriorates. This notion is consistent with a staging paradigm proposed for T2D, rather than a categorial differentiation between stages [2]. Consistent with this, the follow-up T2D signature was enriched for pathways central to metabolic inflexibility—lipid and fatty-acid metabolism and branched-chain amino-acid degradation—while apoptosis and autophagy-related processes were comparatively reduced, suggesting coordinated remodeling of nutrient handling and cellular stress responses with advancing disease [3, 20]. Notably, 111 of the 185 differentially expressed proteins were not reported in prior circulating T2D proteomics comparisons, extending earlier cross-sectional work that typically assessed a more limited set of proteins and also a less structured follow up period [8, 21].

At the individual level, within-person changes in plasma protein abundance (Δ, baseline to follow-up) associated with changes in key clinical features, including fasting- and 2-hour glucose, HbA1c, insulin sensitivity/resistance, β-cell function and IHL content. The change in IGSF9 showed the strongest positive association with HbA1c and an inverse association with insulin sensitivity, and changes in PVR were associated with multiple T2D-related traits (including fatty liver index, OGIS, fasting and 2-hour glucose, and disposition index) specifically in the longitudinal setting, nominating these proteins as dynamic correlates of metabolic deterioration.

Although proteomic profiles at baseline did not allow for a prediction of the outcomes, we identified stage-associated signatures with potential value for risk stratification in prediabetes, a clinically important goal given that only a subset of individuals progress to T2D and preventive resources should be prioritized for those at highest risk [22, 23]. Among 36 proteins tested at baseline, SELE, IGSF9, and PRCP were significantly associated with incident T2D. Baseline association analyses also highlighted IL18R1 as having the strongest adverse association with HbA1c, aligning with its role in interleukin-18 signaling and prior links to metabolic deterioration [24]. Glucagon showed baseline-only associations with insulin sensitivity, IHL content, and VAT volume, consistent with early dysregulation of glucagon-related biology in prediabetes [25, 26].

Genome-wide pQTL mapping in prediabetes at follow-up identified 555 independent loci associated with protein abundance (398 *cis* and 157 *trans*), including 37 novel *trans*-signals when compared with prior pQTL studies, indicating substantial overlap with established circulating protein pQTLs while capturing additional prediabetes-relevant regulation. Condition-aware analyses suggested that genetic regulation can be stage-dependent: mash analysis across 2,460 *cis*-pQTLs identified 86 variants with differential effects in prediabetes relative to T2D, exemplified by rs6107718 (associated with lower plasma CHGB specifically in prediabetes) and differential *cis*-signals for SELE, SCLY, and ACP5. These proteins have been previously linked to inflammatory response, oxidative stress and metabolic disease [27–29]. In parallel, 700 *cis*-pQTLs showed concordant effects across both prediabetes and T2D, exemplified by rs28636067 associated with lower plasma PDCD6 in both conditions, while a subset displayed opposite effect directions between stages, consistent with shifting regulatory context during progression. Among shared genetically regulated plasma proteins, those also differentially expressed between prediabetes and T2D were enriched for IL-8 regulation/production, T-cell–mediated immunity, and sterol metabolism, linking inherited regulation to immune and metabolic pathways implicated in T2D, while noting the immune orientation of the proteomics panel [30].

Finally, mediation analyses provided evidence compatible with protein abundance acting as an intermediate between genotype and clinical traits. Differential pQTL–protein pairs tested across 11 phenotypes yielded significant SNP–protein–phenotype triplets involving 29 plasma proteins, with prominent effects on body fat distribution and insulin sensitivity—phenotypes repeatedly coupled to proteomic variation during prediabetes and important determinants in the pathophysiology of T2D. Several proteins were shown to be important to liver fat. CHGB, a matrix protein expressed in liver tissue (GTEx), showed the strongest adverse effect on liver fat despite no prior link to hepatic function, highlighting it as a promising target for future studies [31, 32]. Galectin-1 exhibited the strongest protective effect on liver fat and is known to regulate cell adhesion, inflammation, signaling, and lipogenesis, consistent with previous links to hepatic steatosis and obesity [33, 34]. SELE, whose baseline levels predicted future T2D and were associated with body fat, liver fat, and insulin resistance in both baseline and Δ-analyses, also significantly mediated these same traits, underscoring its potential as a strong candidate for future investigation. We selected mediation analysis over Mendelian randomization as it has been successfully applied in previous causal studies of circulating proteomics in T2D [11]. Mediation analysis decomposes the total effect into direct and indirect (protein-mediated) components. Because mediation models use the measured protein levels directly, they can remain informative even when the genetic instrument for the protein is weak (in case of modest sample size). In contrast, Mendelian randomization relies entirely on the strength of the genetic instrument and therefore requires large sample sizes or strong pQTLs to obtain stable and unbiased causal estimates [35]. However, mediation analysis does not prove biological causality, which can only be established by follow-up proof of concept experimental studies.

Several considerations support a constructive interpretation and motivate next steps. The number of participants progressing to T2D was smaller than the prediabetes group, which may have reduced sensitivity for detecting weaker or context-specific genetic effects in established T2D; larger datasets will help refine stage-dependent regulation. The study population was largely of European ancestry, providing a strong foundation for discovery but highlighting the importance of replication in more diverse ancestries to enhance generalizability. In addition, the targeted nature of the Olink Explore panel enables robust, scalable measurement but does not capture the full circulating proteome and is enriched for immune-related proteins, warranting caution when interpreting pathway enrichment. However, we captured more than 3000 plasma proteins, which is a robust fraction of the full circulating proteome.

Overall, our results demonstrate how longitudinal plasma proteomics integrated with genetics and deep phenotyping can resolve stage-specific molecular signatures, identify candidate biomarkers for risk stratification and monitoring, and nominate shared and context-dependent regulatory mechanisms that may inform prevention strategies for progression from prediabetes to T2D and potential therapeutic targets.

## Methods

### Study cohort

The Prediabetes Lifestyle Intervention Study (PLIS) is a longitudinal, stratified and randomized multicenter trial conducted in University Hospitals in Germany within the framework of the German Center for Diabetes Research (DZD) to evaluate lifestyle intervention-based strategies for achieving remission to NGR in individuals at risk of T2D [12]. A total of 458 participants were aged 18–75 years with a BMI ≤45 kg/m² and underwent oral glucose tolerance test (OGTT). Inclusion criteria were either elevated fasting glucose (> 100mg/dl < 126mg/dl) and/or elevated 2-hour glucose (> 140 mg/dl < 200 mg/dl) [36]. At baseline and follow-up 6-year, participants underwent deep metabolic phenotyping, including whole body MRI / ^1^H-MRS to determine whole body fat distribution, including VAT volume and IHL content, respectively. The trial was approved by the ethics committee of Eberhard-Karls University of Tübingen, and all participants provided written informed consent.

### Genotyping and imputation

The participants were genotyped using the Illumina GSAMD-24v1 and GSAMD-24v2 chips. The .idat files were genotyped separately per chip version using a custom manifest file and the iaap-cli gencall algorithm. The resulting .gtc files were converted to vcf using *bcftools* with hg19/b37 as the reference genome. These files were converted to *PLINK* (version 2.0) files on which variant and sample quality control (QC) were performed (as described below) [37]. The genotype data was imputed using the Munich imputation server with haplotypes derived from the Human Reference Consortium panel [38]. The imputed genotype data was processed, and QC was performed at the variant level and the sample level using *PLINK*.

### Variant and sample quality control

We performed sample QC by comparing individuals to the multidimensional scaling (MDS) projection of the 1000 Genomes Project reference panel and excluded ancestry outliers by retaining only those clustering to the European reference samples. We also exclude highly related individuals and individuals for whom there was a mismatch between reported sex and genetic sex. Highly heterozygous samples (heterozygosity greater than 3 standard deviations) were also excluded from the analysis.

Variants were filtered out if their missingness was greater than 98% and Hardy-Weinberg equilibrium p-value was smaller than 1 × 10^-5^. Finally, biallelic variants with info score greater than 0.5 were included in our analysis. The final imputed dataset after QC contained 458 individuals and 17,465,456 genetic variants.

### Proteomics data

We performed proximity extension–based proteomics using the Olink Explore panel at baseline and at the 6-year follow-up after lifestyle intervention. Both timepoints were included in the same measurement series and randomized across plates to minimize batch effects. At baseline, proteomic data was available for 2944 proteins measured in 458 individuals. We excluded 405 proteins with >40% sample missingness or with relative expression values below the limit of detection and 3 proteins with bimodal distribution, leaving 2536 proteins across 458 individuals for the baseline analysis.

At the 6-year follow-up, proteomic data was available for 458 individuals. Among these were 49 individuals with NGR, 88 met the criteria for T2D and 321 individuals remained with prediabetes. Of the 2944 proteins assayed, 418 were excluded due to >40% missingness or values below the detection limit, as well as 3 proteins due to their bimodal distribution. After quality control, 2523 proteins and 458 individuals were included in the follow-up analysis.

We included all or a combination of the following covariates in our analyses which had a significant effect on protein levels: age, age^2^, genetic sex, sample storage time (in years), site of sample collection, and mean protein expression per sample (mean NPX).

### Differential protein expression analysis

The relative protein expression measurements were rank normalized and transformed for differential expression analysis. We used a linear mixed model as implemented in the R-package *limma* (version 3.58.1) to determine the differences in protein levels within the same individuals at the two timepoints [39]:

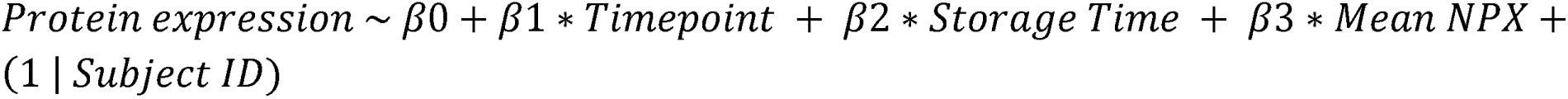

Using *limma,* we analyzed the differences in the protein levels between the groups at the two time points:

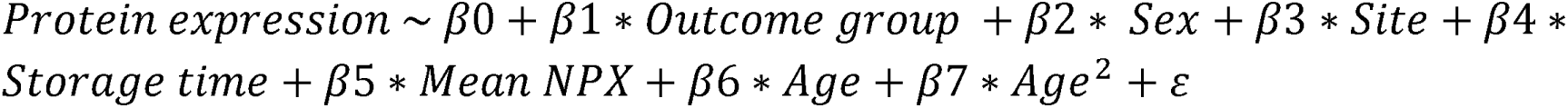

After fitting the model, proteins with log_2_ fold change greater than 0 and FDR < 0.05 were determined to be upregulated while those with log_2_ fold change smaller than 0 and FDR < 0.05 were defined as downregulated proteins.

Gene set enrichment analysis (GSEA) was performed to identify perturbed biological pathways using the *clusterProfiler* R-package (version 4.10.1) for *Homo sapiens*, based on annotations from the Gene Ontology (GO) and KEGG databases [40]. All proteins were ranked by their log_2_ fold change to perform the analysis.

### ΔPlasma protein expression analysis

For the plasma proteins identified as differentially expressed proteins between the prediabetes and the T2D groups at follow-up, we calculated the Δprotein expression (follow up - baseline). In a linear regression framework, the Δprotein expression values were associated with T2D or prediabetes status as follows:

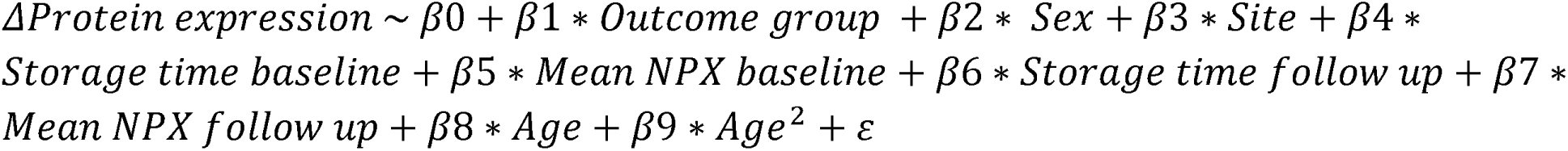

Proteins that were identified as significantly associated (FDR < 0.05) with T2D status were further analyzed. For these proteins, their baseline expression was associated with the baseline value of 11 clinical features. Furthermore, for the same proteins, their log_2_ fold change (log (follow up) - log(baseline)) was associated with log_2_ fold change of the 11 clinical features. The features assessed include body fat percentage, visceral adipose tissue volume, intra-hepatic lipid content, disposition index, fatty-liver index, Homeostatic Model Assessment for Insulin Resistance (HOMA-Insulin Resistance), insulinogenic index, Oral Glucose Insulin Sensitivity (OGIS), fasting glucose, 2-hour glucose and HbA1c.

### Baseline plasma protein levels as predictors of T2D status

Plasma proteins whose expression changes were significantly associated with T2D status were further evaluated by testing whether their baseline expression levels predicted T2D status at follow-up using logistic regression adjusted for age, sex, storage time and mean NPX / sample. The regression coefficients were exponentiated to obtain odds ratios (ORs), which indicate the relative change in odds of developing T2D per standard deviation.

### Genome-wide pQTL analysis

After filtering variants based on the QC criteria described above, we calculated a genetic relationship matrix (GRM) using *GCTA* (version 1.93.0) [41]. The GRM was included in the mixed model to account for relatedness and population structure. Using the genetic variant data and genetic relationship matrix, we performed association using the MLMA linear mixed model algorithm of *GCTA* to identify pQTL. The pQTL analysis presented in our study was performed in two groups:

- 88 individuals with T2D at follow-up
- 321 individuals with prediabetes at follow-up

Note that protein expression data were adjusted for covariates, and the residuals were inverse-normal transformed for the pQTL analysis.

pQTL and further analysis was restricted to genetic variants with minor allele frequency (MAF) > 5%. The significance for pQTLs was defined as the genome-wide significance p-value < 5×10^-8^. To determine independent signals, we used *GCTA-COJO*, which applies an approximate conditional and joint stepwise model selection procedure at each protein-associated genetic locus. We defined *cis-*pQTLs as variants which are located within 1Mb upstream or downstream of the transcription start site of the gene encoding for that protein. Any pQTL located outside of the 1Mb fragment on the same chromosome or a different chromosome was defined as a *trans-*pQTL. All further investigations were restricted to *cis*-pQTLs.

To determine the novelty of pQTLs associated with plasma protein levels in individuals with prediabetes at follow-up, we compared our findings with results from 47 previously published studies of circulating proteome pQTLs (Supplementary table 5). For each study, we collected genome-wide significant summary statistics and metadata, including the author, PubMed ID, discovery cohort size, genomic coordinates of lead variants, UniProt identifiers, effect and non-effect alleles, allele frequencies, effect sizes and directions, mapped genes, p-values, and cis/trans classification. Genomic coordinates were standardized across studies using the *liftOver* function from the R-package *rtracklayer*. To assess novelty, we first matched proteins by name or gene symbol, then identified overlapping variants between previously reported lead SNPs and our pQTLs within a ±1 Mb genomic window. A pQTL was considered novel if no previously reported genome-wide significant association was found within this window for the corresponding protein.

### Differential pQTL analysis

To identify genetic variants with condition-specific effects or shared effects between prediabetes and T2D, we used a mixture model framework as implemented in the R-package *mashr* (version 0.2.79) to perform differential pQTL analysis [42]. This analysis was conducted only for *cis*-pQTLs as they have higher statistical power compared to *trans*-genetic effects [32, 43]. We analyzed 2460 proteins with *cis*-pQTLs between the prediabetes and T2D groups. Note that we excluded 66 proteins encoded by X-chromosome genes, as association testing was not performed on the X-chromosome.

From the data we extracted a subset containing strong signals, i.e. SNP with the most significant association p-value in the *cis*-locus, to compute model priors. We also extracted random signals from the data to learn the null distribution as well as to fit the mash mixture model. The data format was changed to HDF5 format for easier processing in R. This was achieved using the data analysis pipeline in the Jupyter notebook: https://github.com/stephenslab/gtexresults/blob/master/workflows/fastqtl_to_mash.ipynb. Using the *mashr* package in R, we fit the mash model to estimate the mixture proportions followed by computing the posterior effect estimates as well as the local false sign rate (LFSR). In our analysis, we defined differential pQTL as those with LFSR < 0.05 in only one of the conditions.

For proteins corresponding to shared pQTLs that were also significantly differentially expressed between the T2D and prediabetes groups at follow-up, we performed an over-representation analysis to identify enriched biological processes using the R-package *clusterProfiler*. The analysis was conducted for *Homo Sapiens* using the Gene Ontology (GO) database, with all GO-annotated genes used as background set.

### Mediation analysis

Our final objective was to identify causal relationships between plasma proteins and T2D-relevant clinical traits. To this end, we performed mediation analysis using the R-package *mediation* (version 4.5.1). For each pQTL–protein–phenotype triplet, the framework estimates two effects: (1) the effect of the genetic variant (pQTL) on protein levels, and (2) the effect of the protein on the clinical trait, after accounting for the direct effect of the genetic variant. Biologically, this approach tests whether the genetic variant influences the phenotype indirectly through its effect on the protein, implying a causal mediating role of the protein.

For each relationship, linear models were fitted for the mediator (protein ∼ SNP + covariates) and outcome (phenotype ∼ SNP + protein + covariates), adjusting for age, age², sex, collection center, storage time, and mean NPX per sample. The average causal mediation effect (ACME) was estimated using 1,000 nonparametric bootstrap simulations, and multiple testing was controlled using the FDR Benjamini–Hochberg procedure. We report FDR-adjusted p-values and ACME values, representing the per-allele mediated effect (in SD units) of the pQTL on the phenotype via the protein.

## Data Availability

All data produced in the present study are available upon reasonable request to the authors.

https://github.com/hmgu-itg/PLIS-Project

## Acknowledgements

Archit Singh, Dr Mauro Tutino and Dr Ozvan Bocher have received funding from the European Union’s Horizon 2020 research and innovation program under Grant Agreement No 101017802 (OPTOMICS). We gratefully acknowledge the contribution of the PLIS cohort participants as well as the technical and nursing staff contributing greatly to data collection. PLIS and this post hoc analysis were supported by the German Center for Diabetes Research, which is funded by the German Federal Ministry for Education and Research and the German states where its partner institutions are located (01GI0925). We acknowledge the support of the Core Facility-Metabolomics and Proteomics and the Core Facility Genomics at Helmholtz Munich for processing the samples and generating the data.

---

Supplementary Figure 1: CONSORT flow chart describing the total number of individuals available for analysis at baseline and those that have follow-up data available in each outcome group. NGR = Normal glucose regulation at follow-up, T2D = Type 2 diabetes.

Supplementary Figure 2: Volcano plots showing the comparison of plasma protein levels between groups at baseline.

A. Normal glucose regulation (NGR) group versus Prediabetes group

B. T2D group versus Prediabetes group

Supplementary Figure 3: Volcano plots showing the comparison of plasma protein levels within groups at baseline versus follow-up. Dots with red color show upregulated proteins and those with blue show downregulated proteins at follow-up.

A. Normal glucose regulation (NGR) group

B. Prediabetes group

C. T2D group

Supplementary Figure 4: Volcano plot showing the comparison of plasma protein levels in individuals with normal glucose regulation (NGR) versus the prediabetes group at follow-up.

Supplementary Figure 5: Forest plot showing the posterior effect sizes of the shared pQTLs with opposite effects in the prediabetes versus the T2D conditions. The y-axis shows the plasma proteins and the corresponding shared pQTLs that showed opposite effects between prediabetes and T2D conditions at follow-up. T2D = Type 2 diabetes

Supplementary Table 1: A table showing the baseline characteristics of PLIS participants included in the analysis (all diagnosed with prediabetes). BMI = Body Mass Index, VAT [L] = Visceral Adipose Tissue [Liters], IHL = Intrahepatic Lipids, FLI = Fatty Liver Index, HOMA-IR = Homeostasis Model Assessment for Insulin Resistance, OGIS = Oral Glucose Insulin Sensitivity Note that Disposition index measures the pancreatic β-cell function and the Insulinogenic index measures how effectively the pancreas release insulin in the early phase after eating.

Supplementary Table 2: A table showing the characteristics of PLIS cohort participants included in the analysis. Within group comparisons of anthropometric and clinical traits have been shown with significant changes highlighted in bold. NGR = Normal Glucose Regulation, PreD = Prediabetes, T2D = Type 2 Diabetes at follow-up. BMI = Body Mass Index, VAT [L] = Visceral Adipose Tissue [Liters], IHL = Intrahepatic Lipids, FLI = Fatty Liver Index, HOMA-IR = Homeostasis Model Assessment for Insulin Resistance, OGIS = Oral Glucose Insulin Sensitivity Note that Disposition index measures the pancreatic β-cell function and the Insulinogenic index measures how effectively the pancreas releases insulin in the early phase after eating.

Supplementary Table 3: Table of plasma proteins showing significant differential expression between the T2D and the prediabetes groups at follow-up.

Supplementary Table 4: The list of circulating proteome studies related to T2D which were analyzed to assess the novelty of our results.

Supplementary Table 5: Table showing the list of clinical features, plasma proteins and their association cross-sectionally at baseline and longitudinally over time. VAT_l = Visceral Adipose Tissue [Liters], IHL = Intrahepatic Lipids, DI = Disposition index, FLI = Fatty liver index, HOMA-IR = Homeostasis Model Assessment for Insulin Resistance, IGI = Insulinogenic index, OGIS = Oral Glucose Insulin Sensitivity

Supplementary Table 6: Table of independent pQTLs in the prediabetes group at follow-up.

Supplementary Table 7: The list of studies used to investigate the novelty of the identified independent pQTLs reported in Supplementary Table 6.

Supplementary Table 8: Table of differential pQTLs in the prediabetes group compared to the T2D group at follow-up.

Supplementary Table 9: Table of shared pQTLs between the prediabetes and the T2D groups at follow-up.

Supplementary Table 10: Table listing the pQTLs, the proteins and the clinical features as well as the ACME and the FDR-corrected p-values from the mediation analysis. VAT_l = Visceral Adipose Tissue [Liters], FLI = Fatty liver index, HOMA-IR = Homeostasis Model Assessment for Insulin Resistance, IGI = Insulinogenic index, OGIS = Oral Glucose Insulin Sensitivity

