## Supplementary Figure 1 for "Longitudinal Proteogenomic Analysis Reveals Mechanistic Insights into the Progression from Prediabetes to Type 2 Diabetes"

### Slide 1
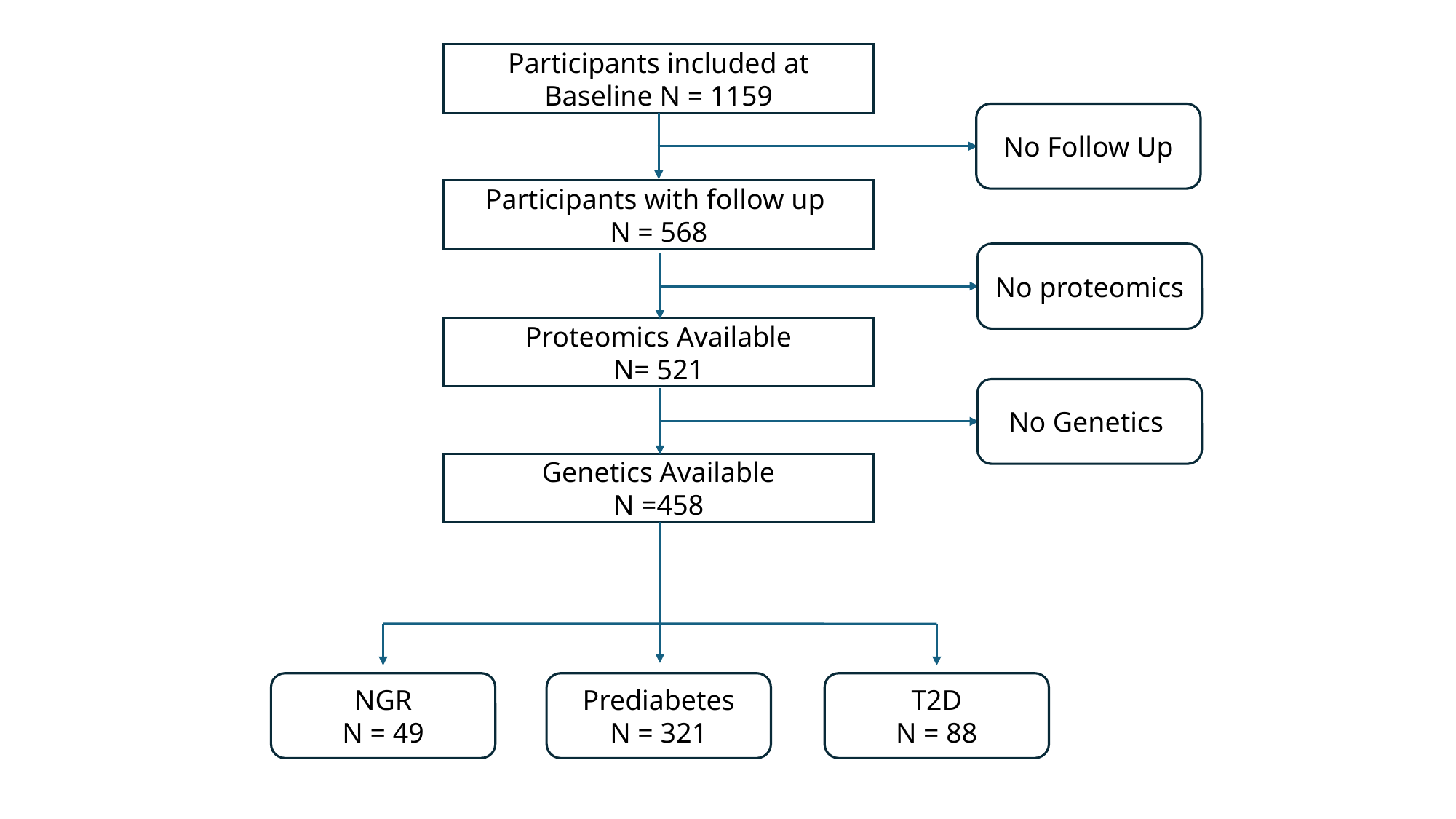

Participants included at Baseline N = 1159
No Follow Up
Participants with follow up
N = 568
No proteomics
Proteomics Available
N= 521
No Genetics
Genetics Available
N =458
NGR
 N = 49
Prediabetes
N = 321
T2D
N = 88
