## Supplementary figures and images for "Longitudinal Proteogenomic Analysis Reveals Mechanistic Insights into the Progression from Prediabetes to Type 2 Diabetes"

### Supplementary Figure 2

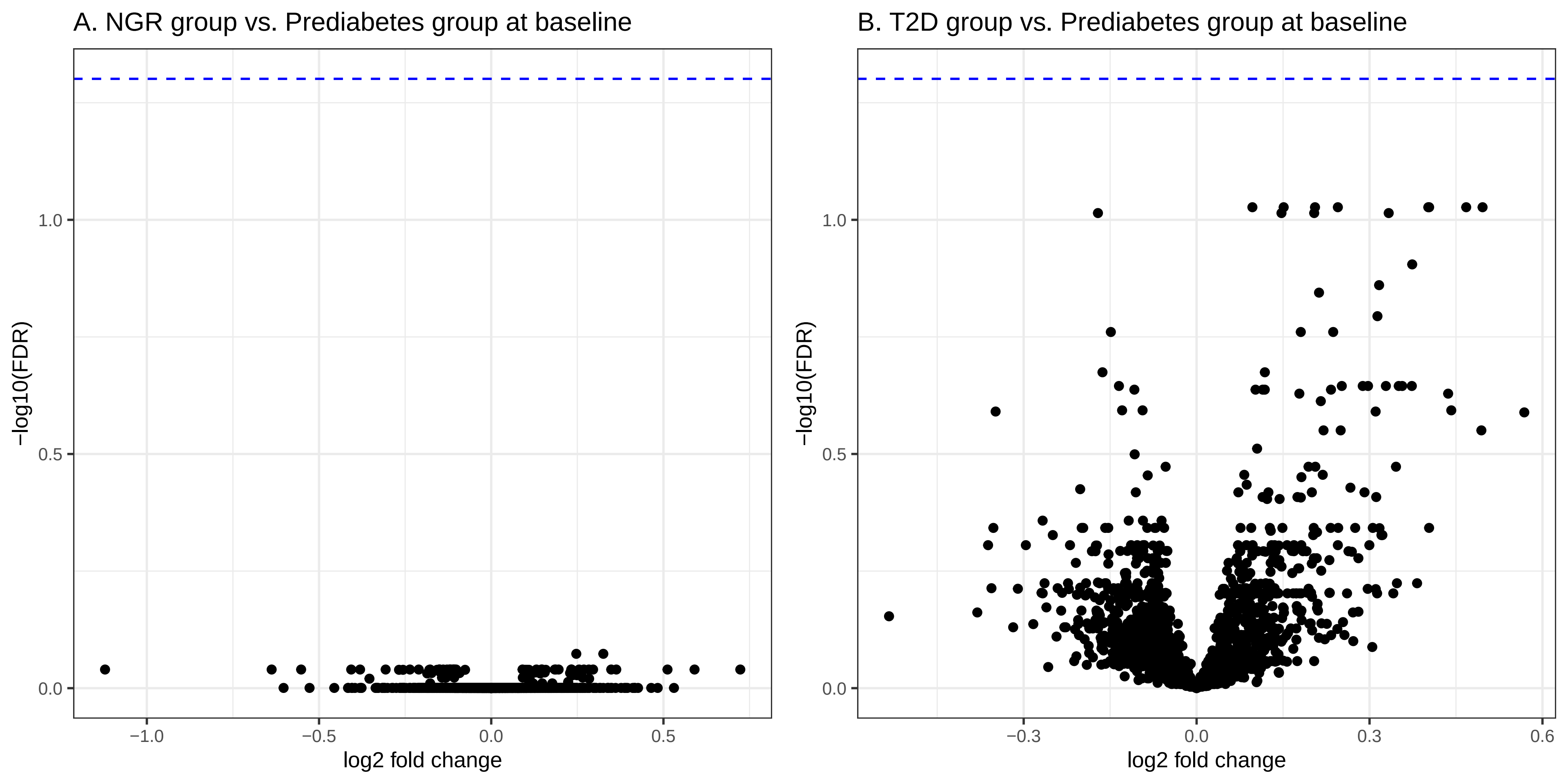

### Supplementary Figure 3

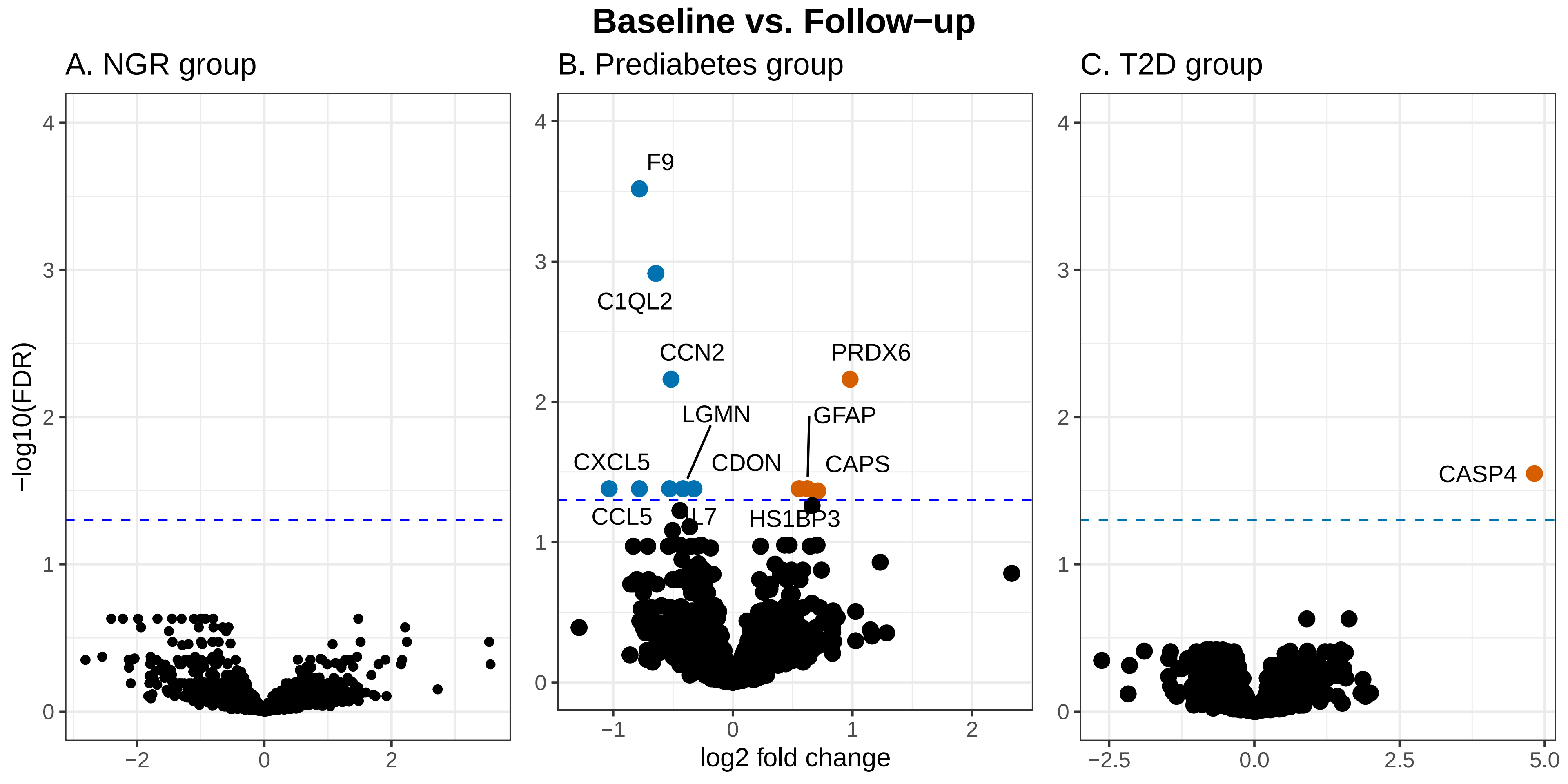

### Supplementary Figure 4

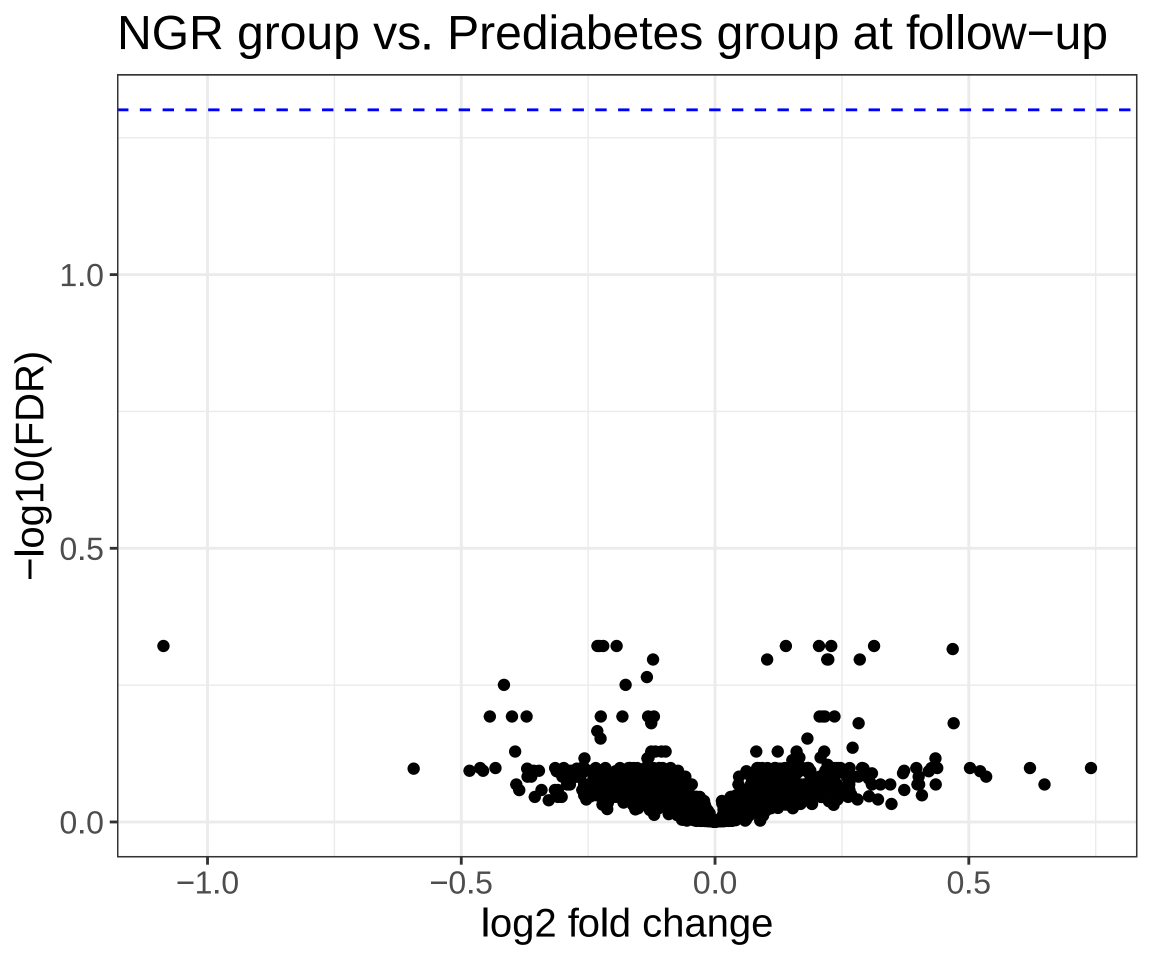

### Supplementary Figure 5

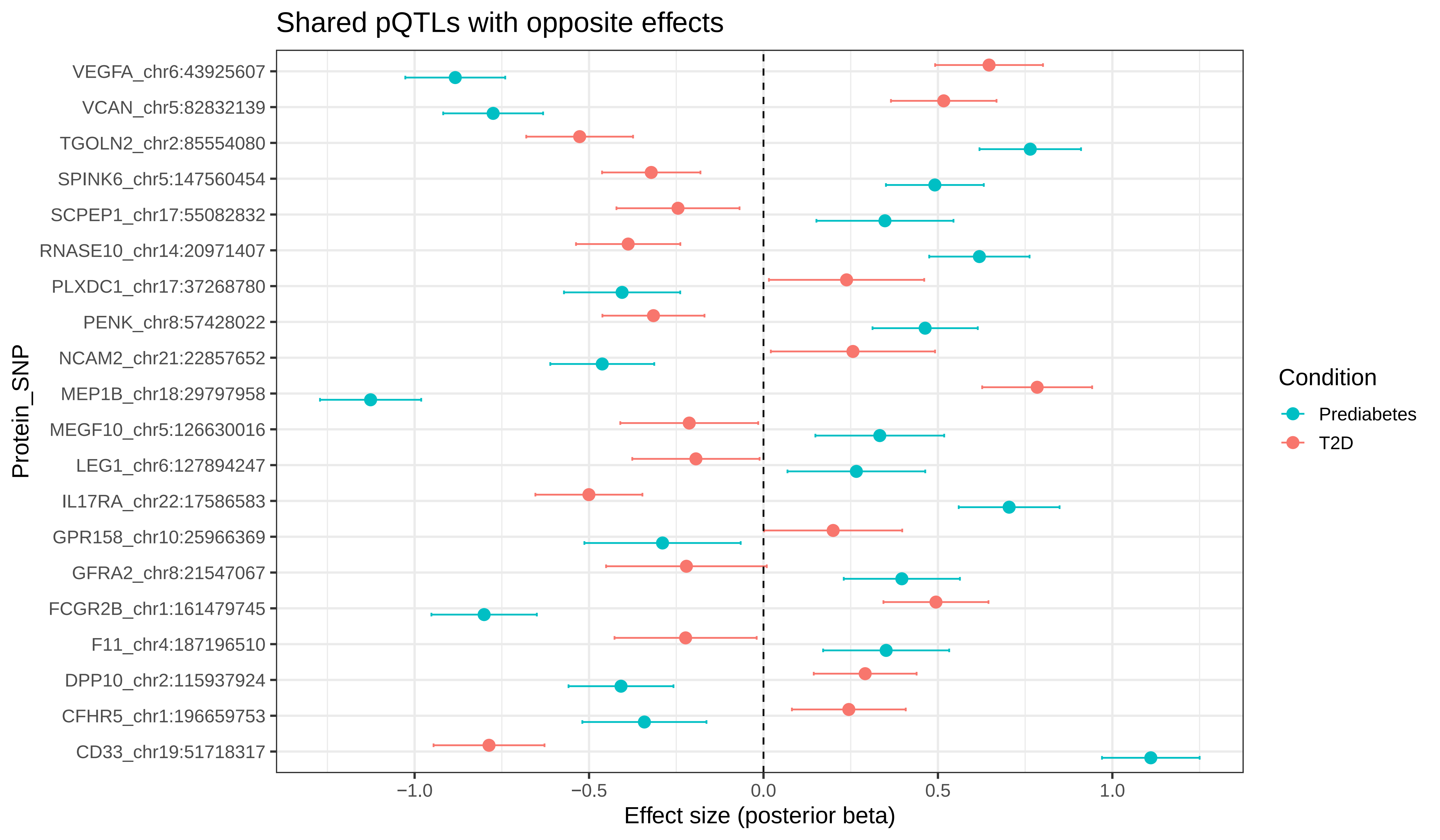
