## Supplementary Table 1 for "Longitudinal Proteogenomic Analysis Reveals Mechanistic Insights into the Progression from Prediabetes to Type 2 Diabetes"

| Variable | N = 458[1] |
|---|---|
| **Sex** | |
| Female | 247 (54%) |
| Male | 211 (46%) |
| **Age** | 61 [54-67] |
| **BMI** | 29.2 [26.1-33.0] |
| **Body Fat [%]** | 33 [26-42] |
| **VAT [L]** | 4.91 [3.22-6.68] |
| **IHL [%]** | 6 [3-11] |
| **FLI** | 65 [38-85] |
| **Disposition Index** | 1.99 [1.28-3.14] |
| **HOMA-IR** | 3.04 [2.16-4.42] |
| **Insulinogenic Index** | 0.78 [0.51-1.48] |
| **OGIS** | 323 [281-366] |
| **Glucose 0 [mg/dL]** | 106 [101-112] |
| **Glucose 2h [mg/dL]** | 135 [112-156] |
| **HbA1c [%]** | 5.80 [5.60-6.00] |

[1] n (%); Median [Q1-Q3]
