## Supplementary Table 2 for "Longitudinal Proteogenomic Analysis Reveals Mechanistic Insights into the Progression from Prediabetes to Type 2 Diabetes"

| Variable | NGR Baseline N = 49[1] | NGR Follow-up N = 49[1] | NGR p-value[2] | PreD Baseline N = 321[1] | PreD Follow-up N = 321[1] | PreD p-value[2] | T2D Baseline N = 88[1] | T2D Follow-up N = 88[1] | T2D p-value[2] |
|---|---|---|---|---|---|---|---|---|---|
| **Age** | 53 [46-64] | 60 [52-71] | **<0.001** | 62 [55-68] | 68 [62-74] | **<0.001** | 62 [54-67] | 68 [60-73] | **<0.001** |
| **BMI** | 29.9 [26.9-33.6] | 27.6 [25.9-31.9] | **<0.001** | 28.7 [25.8-32.3] | 28.3 [25.3-32.1] | **<0.001** | 30.1 [27.2-33.9] | 30.5 [27.3-33.8] | **0.027** |
| **Body Fat [%]** | 34 [24-40] | 32 [22-38] | **0.002** | 33 [26-43] | 31 [24-41] | **<0.001** | 35 [27-43] | 37 [26-44] | 0.5 |
| **VAT [L]** | 4.16 [2.52-7.67] | 3.57 [2.17-6.02] | **0.037** | 4.90 [3.18-6.62] | 4.76 [3.31-6.48] | **0.005** | 5.05 [3.83-6.76] | 5.76 [4.43-6.76] | **0.005** |
| **IHL [%]** | 5.2 [1.6-9.3] | 2.0 [1.1-3.4] | **0.008** | 5 [2-11] | 4 [2-10] | **0.017** | 7 [5-13] | 12 [4-15] | 0.3 |
| **FLI** | 65 [34-89] | 41 [18-76] | **<0.001** | 63 [35-83] | 54 [26-79] | **<0.001** | 73 [52-89] | 79 [54-91] | 0.066 |
| **Disposition Index** | 2.5 [1.6-3.8] | 2.9 [2.2-4.8] | **0.030** | 2.14 [1.38-3.27] | 2.09 [1.39-3.18] | 0.4 | 1.24 [0.90-2.08] | 0.97 [0.68-1.35] | **<0.001** |
| **HOMA-IR** | 2.79 [2.22-3.95] | 2.34 [1.82-3.30] | **0.012** | 3.03 [2.08-4.25] | 3.22 [2.24-4.71] | **0.045** | 3.4 [2.3-5.1] | 5.2 [3.3-9.0] | **<0.001** |
| **Insulinogenic Index** | 0.95 [0.58-1.61] | 0.96 [0.60-1.54] | 0.081 | 0.81 [0.52-1.51] | 0.81 [0.51-1.26] | **0.007** | 0.62 [0.39-1.23] | 0.57 [0.30-0.89] | 0.3 |
| **OGIS** | 339 [288-388] | 398 [366-439] | **<0.001** | 325 [283-367] | 339 [306-374] | **0.003** | 299 [259-346] | 285 [238-324] | **<0.001** |
| **Glucose 0 [mg/dL]** | 102 [100-106] | 92 [88-97] | **<0.001** | 105 [101-111] | 105 [99-111] | **0.016** | 113 [106-119] | 128 [117-136] | **<0.001** |
| **Glucose 2h [mg/dL]** | 123 [102-151] | 103 [86-117] | **<0.001** | 132 [110-153] | 127 [106-147] | **0.044** | 151 [132-175] | 214 [183-237] | **<0.001** |
| **HbA1c [%]** | 5.50 [5.30-5.60] | 5.40 [5.20-5.50] | **0.046** | 5.80 [5.60-6.00] | 5.80 [5.60-6.00] | **0.030** | 5.90 [5.70-6.10] | 6.30 [6.00-6.60] | **<0.001** |

[1] Median [Q1-Q3]

[2] Paired Wilcoxon test per subject
